# Assessing the risk of early-onset dementia within 5 years of cancer diagnosis

**DOI:** 10.64898/2026.02.12.26346204

**Authors:** Corinne E. Joshu, Maylin Palatino, Xiaoqiang Xu, Yiyi Zhou, Eryka Saylor, Jacqueline E. Rudolph, Karine Yenokyan, Keri Calkins, Bryan Lau

**Author notes:** Funding: Research reported in this publication was funded in part by the National Cancer Institute of the National Institutes of Health under Award Number R01CA250851. The content is solely the responsibility of the authors and does not necessarily represent the official views of the National Institutes of Health. Data Availability: The data that support the findings of this study are under the authority of the Centers for Medicare & Medicaid Services (CMS) and administered by ResDAC. Investigators may reuse these data if they independently meet CMS requirements and obtain both CMS reuse approval and permission from the study’s NIH program officer.

## Abstract

**Objective:** To evaluate risk of early-onset dementia (EOD) after diagnosis of cancer among Medicaid beneficiaries.

**Design:** Longitudinal observational study of Medicaid enrollment, inpatient, and outpatient claims data from 26 states and Washington, DC, 2001-2019.

**Methods:** Beneficiaries aged 18-64 with ≥6 months of enrollment were matched 1:1 on cancer status (lung, colon, breast, prostate) by age, sex, race, year and state. We estimated the weighted cumulative incidence functions of EOD at 1, 2, and 5 years after cancer diagnosis using the Aalen-Johansen estimator to account for the competing risk of death and cluster stratified analyses to account for matching. We calculated the corresponding risk differences (RD) and 95% confidence intervals (CI) using the 2.5^th^ and 97.5^th^ percentile of point estimates from 500 bootstrap resamples.

**Results:** The 5-year risk of EOD was 4.7% (95%CI: 4.5,5.0) and 4.7% (95%CI: 4.4, 4.9) among those with and without lung cancer, respectively (RD:0.08; 95%CI: -0.27,0.42). The 5-year risk of EOD was 4.1% (95%CI: 3.8, 4.4) and 3.9% (95%CI:3.7,4.3) among those with and without colon cancer, respectively, (RD 0.18; 95%CI: -0.25,0.55). The 5-year risk of EOD was 3.0% (95%CI: 2.8,3.1) and 2.9% (95%CI: 2.7,3.0) among those with and without breast cancer, respectively, (RD 0.10; 95%CI: -0.14,0.43). The 5-year risk of EOD was 4.6% (95%CI: 4.3,4.9) and 5.3% (95%CI: 4.9,5.7) among those with and without prostate cancer, respectively; those with prostate cancer had a lower EOD risk (RD -0.66; 95%CI: -1.2,-0.16).

**Conclusions:** EOD incidence peaked at 4-5% among beneficiaries with and without cancer. Diagnosis of lung, colon, breast and prostate cancers were not strongly associated with EOD within 5 years. Additional work is needed to identify risk factors for EOD.

## Introduction

Early-onset dementia (EOD), which is commonly defined as dementia occurring before the age of 65 (1), has been increasing globally and accounts for approximately 13% of all dementias (2, 3). An aging population and improved identification and diagnosis of EOD likely both contribute to the increase over time. EOD often impacts people during years of increased occupational and familial responsibilities creating unique challenges (2-4). In addition, EOD may have more aggressive progression and atypical features (2, 3). A growing body of research has sought to identify non-genetic risk factors for EOD by examining lifestyle factors and comorbid conditions associated with average onset dementia (5-8).

Cancer is a significant co-morbidity. While most cancer occurs at older ages, the incidence of several cancers has been increasing at younger ages (9, 10). Meta-analyses of prior population-based studies have reported modest but consistent inverse associations between cancer and dementia, but have highlighted the challenges in accounting for biases, including competing risks and survival bias, in these findings (11, 12). Site-specific findings for common cancers including lung, colorectal, breast, and prostate were heterogenous but generally null or inverse. Whether these associations are similar for EOD and cancer diagnosed at younger ages has not been well-described. Given the increasing burden of both dementia and cancer at younger ages, more work is needed to characterize the association between common cancers and EOD.

Thus, we evaluated the incidence of EOD among Medicaid beneficiaries with and without cancer who were enrolled in Medicaid between 2000 and 2019 in Washington, DC or one of 26 states. Briefly, Medicaid is a joint federal and state program that provides health coverage to individuals that meet certain criteria including those who are low-income. Our analysis included more than 30,000 cases per cancer type among a population with high co-morbidity burden who thus may be at higher risk of EOD. We addressed concerns of prior study designs by matching beneficiaries with cancer to those without cancer and estimating dementia incidence at 1-, 2-, and 5-years post-date of cancer diagnosis while accounting for the competing risk of death.

## Methods

### Study Sample

We used enrollment, inpatient, and outpatient claims data from the Centers for Medicare and Medicaid Services for Medicaid beneficiaries between 2001 and 2019 in Washinton, DC or one of 26 states (Alabama, Arkansas, Arizona, California, Colorado, Florida, Georgia, Illinois, Indiana, Kentucky, Maryland, Massachusetts, Michigan, Missouri, Mississippi, Nevada, New Jersey, New York, North Carolina, Ohio, Oklahoma, Pennsylvania, South California, Tennessee, Texas, and Washington). The analysis was restricted to beneficiaries 18-64 years old with full benefits, no dual enrollment in Medicare or private insurance, and ≥6 months of enrollment. These criteria allow for complete capture of medical claims, equivalent opportunity for services, and enough enrollment time to identify prevalent conditions. Because incident, not prevalent, cancer was our exposure of interest, beneficiaries with any evidence of a cancer diagnosis identified via ICD-9 or ICD-10 codes on any claim within the first six months of enrollment were excluded (Table S1.) The Johns Hopkins Bloomberg School of Public Health Institutional Review Board determined that this secondary analysis of existing Medicaid claims data meets the criteria for exemption.

### Exposure

We defined incident cancer as having one inpatient claim or two outpatient claims with a cancer-related ICD-9 or ICD-10 code within 2 years; the first service date was used as date of diagnosis (Table S1).(13) We identified incident diagnoses of lung, colon, breast, and prostate cancers.

### Outcome

We defined incident dementia as having one inpatient claim or two outpatient claims with a dementia-related ICD-9 or ICD-10 code within 2 years; the first service date was used at date of diagnosis (S1).(13) Because incident, not prevalent, dementia after cancer diagnosis was our outcome of interest, beneficiaries with any evidence of a dementia diagnosis on any claim prior to the index date (see Matching) were excluded from analysis.

### Covariates

We obtained demographic and enrollment information from the personal summary file, including sex, race/ethnicity (non-Hispanic White, non-Hispanic Black, Hispanic, other), state, and dates of birth, death, enrollment to Medicaid, and disenrollment. We defined Charlson comorbidity conditions -- myocardial infarction (MI), congestive heart failure (CHF), peripheral vascular disease (PVD), cerebrovascular disease (CVD), chronic pulmonary disease (COPD), rheumatic disease, peptic ulcer disease, liver disease, hemiplegia or paraplegia, renal disease, human immunodeficiency virus (HIV), and diabetes--as having one ICD-9 or ICD-10 diagnosis code from the inpatient, outpatient, or long-term care files prior to the index date (Table S1).

### Matching

We created 1:1 sets matched on cancer status for each cancer type of interest using incidence density matching with replacement (Supplementary Methods). Briefly, each beneficiary with cancer was matched to a beneficiary who was cancer-free at the date of diagnosis (index date). Beneficiaries were matched by age within 2 years, sex, race, US state, and enrollment year within 2 years. When more than one beneficiary met the matching criteria, a match was randomly selected from the eligible set. Beneficiaries were eligible to serve as a control for more than one case and beneficiaries with cancer were eligible to serve as a control prior to their date of diagnosis. The index date served as the analytic baseline for each matched pair. We conducted two sensitivity analyses. First, to better account for differences in smoking status among matched pairs for lung cancer, cancer-free matches were selected from beneficiaries with a diagnosis of COPD. Second, to better account for differences in metabolic and cardiovascular health among matched pairs for colon cancer, cancer-free matches were selected from beneficiaries with a diagnosis of any Charlson comorbidity.

### Statistical Analysis

We calculated means and proportions of baseline characteristics of beneficiaries and their matches for each cancer category. Beneficiaries were followed from date of analytic baseline until dementia diagnosis, death, 65^th^ birthday, disenrollment, or 12/31/19, whichever came first. We estimated the crude and weighted cumulative incidence functions (CIFs) of dementia among those with and without cancer using the Aalen-Johansen estimator to account for the competing risk of death and matching, employing robust variance estimation to account for matching. Weighted CIFs were estimated using the estimated and combined exposure (cancer) odds weights and informative censoring weights. The exposure odds weight included age at enrollment (specified using natural cubic splines), baseline diagnosis of MI, CHF, PVD, CVD, diabetes, and HIV, and the number of Charlson comorbidities (0, 1, or ≥2) at baseline as covariates. The censoring weight per beneficiary per record was computed as a ratio of two probabilities: 1) probability of not dropping out with only the time-updated age (specified as natural cubic splines) as the covariate, and 2) the probability of not dropping out with exposure status and time-varying Charlson comorbidity category (within current interval, lag 3 months, and lag 6 months) as additional covariates (14). Both probabilities were obtained using pooled logistic regression. We computed the cumulative censoring weight per beneficiary per record by multiplying the censoring weights from the first through the last record and combined the exposure and censoring weights by multiplying the exposure weight to each record-specific cumulative exposure weight. The distributions of the combined weights were assessed and truncated at the 97.5^th^ percentile. We calculated the crude and weighted CIF at 1, 2, and 5 years after analytic baseline for beneficiaries with and without cancer and the corresponding risk differences (RD). The 95% confidence intervals of the CIFs and RDs were the 2.5^th^ and 97.5^th^ percentile of the point estimates from 500 bootstrap resamples. Analyses were also stratified by age at index date (18-49; 50-64 years), sex, and race/ethnicity. Weighted risk of dementia and RD estimates are described in main text; crude estimates and weighted risk of death, RD estimates and their 95% CI are provided in the supplement.

## Results

Characteristics between beneficiaries with and without cancer were similar due to matching (Table 1). For all cancers, beneficiaries were more likely to be non-Hispanic White than other race/ethnicity; however, prostate cancer had a higher proportion of non-Hispanic Black beneficiaries than any other cancer type. California and New York had the highest proportion of beneficiaries.

**Table 1.** Demographic characteristics, dementia incidence, and median follow-up time of matched Medicaid beneficiaries by cancer type, 2001-2019.

| Characteristic | Lung cancer |  | Colon cancer |  | Breast cancer |  | Prostate cancer |  |
| --- | --- | --- | --- | --- | --- | --- | --- | --- |
|  | Without | With | Without | With | Without | With | Without | With |
| N | 60600 | 60600 | 34691 | 34691 | 72189 | 72189 | 32649 | 32649 |
| Age at baseline, Median [IQR]<sup>1</sup> | 57.59<br>[52.67, 60.96] | 58.02<br>[53.13, 61.51] | 55.99<br>[49.62, 60.48] | 55.99<br>[49.62, 60.48] | 54.32<br>[47.07, 59.75] | 54.63<br>[47.44, 60.09] | 59.17<br>[55.37, 61.75] | 59.66<br>[55.95, 62.40] |
| Sex, N (%) |  |  |  |  |  |  |  |  |
| Female | 29777 (49.1) | 29777 (49.1) | 18085 (52.1) | 18085 (52.1) | 72189 (100.0) | 72189 (100.0) | 0 (0.0) | 0 (0.0) |
| Male | 30823 (50.9) | 30823 (50.9) | 16606 (47.9) | 16606 (47.9) | 0 (0.0) | 0 (0.0) | 32649 (100.0) | 32649 (100.0) |
| Race/Ethnicity, N (%) |  |  |  |  |  |  |  |  |
| Non-Hispanic White | 33184 (54.8) | 33184 (54.8) | 15463 (44.6) | 15463 (44.6) | 32603 (45.2) | 32603 (45.2) | 11488 (35.2) | 11488 (35.2) |
| Non-Hispanic Black | 15152 (25.0) | 15152 (25.0) | 8525 (24.6) | 8525 (24.6) | 16811 (23.3) | 16811 (23.3) | 12470 (38.2) | 12470 (38.2) |
| Hispanic | 2160 (3.6) | 2160 (3.6) | 2264 (6.5) | 2264 (6.5) | 4921 (6.8) | 4921 (6.8) | 1573 (4.8) | 1573 (4.8) |
| Other | 10104 (16.7) | 10104 (16.7) | 8439 (24.3) | 8439 (24.3) | 17854 (24.7) | 17854 (24.7) | 7118 (21.8) | 7118 (21.8) |
| US State, N (%) |  |  |  |  |  |  |  |  |
| AL | 2058 (3.4) | 2058 (3.4) | 1051 (3.0) | 1051 (3.0) | 1355 (1.9) | 1355 (1.9) | 824 (2.5) | 824 (2.5) |
| AR | 299 (0.5) | 299 (0.5) | 111 (0.3) | 111 (0.3) | 283 (0.4) | 283 (0.4) | 83 (0.3) | 83 (0.3) |
| AZ | 534 (0.9) | 534 (0.9) | 429 (1.2) | 429 (1.2) | 1012 (1.4) | 1012 (1.4) | 419 (1.3) | 419 (1.3) |
| CA | 10756 (17.7) | 10756 (17.7) | 9179 (26.5) | 9179 (26.5) | 18695 (25.9) | 18695 (25.9) | 7710 (23.6) | 7710 (23.6) |
| CO | 389 (0.6) | 389 (0.6) | 217 (0.6) | 217 (0.6) | 552 (0.8) | 552 (0.8) | 171 (0.5) | 171 (0.5) |
| DC | 167 (0.3) | 167 (0.3) | 97 (0.3) | 97 (0.3) | 304 (0.4) | 304 (0.4) | 226 (0.7) | 226 (0.7) |
| FL | 4670 (7.7) | 4670 (7.7) | 2459 (7.1) | 2459 (7.1) | 3648 (5.1) | 3648 (5.1) | 2000 (6.1) | 2000 (6.1) |
| GA | 3062 (5.1) | 3062 (5.1) | 1345 (3.9) | 1345 (3.9) | 2593 (3.6) | 2593 (3.6) | 1430 (4.4) | 1430 (4.4) |
| IL | 5176 (8.5) | 5176 (8.5) | 2902 (8.4) | 2902 (8.4) | 5518 (7.6) | 5518 (7.6) | 2923 (9.0) | 2923 (9.0) |
| IN | 794 (1.3) | 794 (1.3) | 301 (0.9) | 301 (0.9) | 833 (1.2) | 833 (1.2) | 327 (1.0) | 327 (1.0) |
| KY | 1588 (2.6) | 1588 (2.6) | 660 (1.9) | 660 (1.9) | 1314 (1.8) | 1314 (1.8) | 403 (1.2) | 403 (1.2) |
| MA | 2033 (3.4) | 2033 (3.4) | 992 (2.9) | 992 (2.9) | 3036 (4.2) | 3036 (4.2) | 973 (3.0) | 973 (3.0) |
| MD | 1392 (2.3) | 1392 (2.3) | 660 (1.9) | 660 (1.9) | 1658 (2.3) | 1658 (2.3) | 714 (2.2) | 714 (2.2) |
| MI | 1963 (3.2) | 1963 (3.2) | 952 (2.7) | 952 (2.7) | 2253 (3.1) | 2253 (3.1) | 1232 (3.8) | 1232 (3.8) |
| MO | 487 (0.8) | 487 (0.8) | 158 (0.5) | 158 (0.5) | 510 (0.7) | 510 (0.7) | 140 (0.4) | 140 (0.4) |
| MS | 318 (0.5) | 318 (0.5) | 153 (0.4) | 153 (0.4) | 266 (0.4) | 266 (0.4) | 145 (0.4) | 145 (0.4) |
| NC | 2944 (4.9) | 2944 (4.9) | 1166 (3.4) | 1166 (3.4) | 2284 (3.2) | 2284 (3.2) | 1307 (4.0) | 1307 (4.0) |
| NJ | 495 (0.8) | 495 (0.8) | 469 (1.4) | 469 (1.4) | 1269 (1.8) | 1269 (1.8) | 477 (1.5) | 477 (1.5) |
| NV | 231 (0.4) | 231 (0.4) | 183 (0.5) | 183 (0.5) | 399 (0.6) | 399 (0.6) | 161 (0.5) | 161 (0.5) |
| NY | 7864 (13.0) | 7864 (13.0) | 4941 (14.2) | 4941 (14.2) | 10849 (15.0) | 10849 (15.0) | 5286 (16.2) | 5286 (16.2) |
| OH | 3512 (5.8) | 3512 (5.8) | 1505 (4.3) | 1505 (4.3) | 3076 (4.3) | 3076 (4.3) | 1330 (4.1) | 1330 (4.1) |
| OK | 310 (0.5) | 310 (0.5) | 114 (0.3) | 114 (0.3) | 201 (0.3) | 201 (0.3) | 87 (0.3) | 87 (0.3) |
| PA | 3480 (5.7) | 3480 (5.7) | 1551 (4.5) | 1551 (4.5) | 3554 (4.9) | 3554 (4.9) | 1748 (5.4) | 1748 (5.4) |
| SC | 352 (0.6) | 352 (0.6) | 174 (0.5) | 174 (0.5) | 464 (0.6) | 464 (0.6) | 188 (0.6) | 188 (0.6) |
| TN | 831 (1.4) | 831 (1.4) | 320 (0.9) | 320 (0.9) | 897 (1.2) | 897 (1.2) | 235 (0.7) | 235 (0.7) |
| TX | 3227 (5.3) | 3227 (5.3) | 1679 (4.8) | 1679 (4.8) | 2898 (4.0) | 2898 (4.0) | 1320 (4.0) | 1320 (4.0) |
| WA | 1668 (2.8) | 1668 (2.8) | 923 (2.7) | 923 (2.7) | 2468 (3.4) | 2468 (3.4) | 790 (2.4) | 790 (2.4) |
| Enrollment period, N (%) |  |  |  |  |  |  |  |  |
| 2001-2005 | 12630 (20.8) | 12630 (20.8) | 6233 (18.0) | 6233 (18.0) | 15004 (20.8) | 15004 (20.8) | 5178 (15.9) | 5178 (15.9) |
| 2006-2010 | 11835 (19.5) | 11835 (19.5) | 6546 (18.9) | 6546 (18.9) | 11979 (16.6) | 11979 (16.6) | 5464 (16.7) | 5464 (16.7) |
| 2011-2015 | 14654 (24.2) | 14654 (24.2) | 9259 (26.7) | 9259 (26.7) | 18902 (26.2) | 18902 (26.2) | 9038 (27.7) | 9038 (27.7) |
| 2016-2019 | 21481 (35.4) | 21481 (35.4) | 12653 (36.5) | 12653 (36.5) | 26304 (36.4) | 26304 (36.4) | 12969 (39.7) | 12969 (39.7) |
| Incident dementia, N (%) | 1085 (1.8) | 1347 (2.2) | 567 (1.6) | 731 (2.1) | 879 (1.2) | 1060 (1.5) | 623 (1.9) | 694 (2.1) |
| Follow-up years, | 1.90 | 0.66 | 1.96 | 1.40 | 2.01 | 1.87 | 1.76 | 1.51 |

| Characteristic | Lung cancer |  | Colon cancer |  | Breast cancer |  | Prostate cancer |  |
| --- | --- | --- | --- | --- | --- | --- | --- | --- |
|  | Without | With | Without | With | Without | With | Without | With |
| Median [IQR]<sup>1</sup> | [0.84, 3.60] | [0.23, 1.59] | [0.87, 3.66] | [0.55, 2.87] | [0.90, 3.73] | [0.82, 3.47] | [0.78, 3.20] | [0.67, 2.93] |

<sup>1</sup>IQR: Interquartile range

### Lung Cancer

The 1-year risk of dementia was 1.9% (95%CI: 1.8, 2.0) among those with lung cancer as compared to 1.1% (95%CI: 1.0,1.2) among those without lung cancer (Figure 1, Table S3). The 5-year risk of dementia rose to 4.7% (95%CI: 4.5,5.0) and 4.7% (95%CI: 4.4, 4.9) among those with and without lung cancer, respectively. Beneficiaries with lung cancer had a 0.76% (95%CI: 0.62,0.90) higher risk of dementia as compared to those without lung cancer at 1 year; findings were similar at 2 years and attenuated at 5 years (RD 0.08; 95%CI: -0.27,0.42). This pattern was similar among beneficiaries 50 or older, males, females, and when stratified by race/ethnicity groups (Figure 2, Figure 3, Table S3). Among beneficiaries less than 50, risk of dementia was lower among beneficiaries with and without cancer than their older counterparts at each time point (Figure 2, Table S3). The 1-year risk of dementia was 0.41% (95%CI: 0.14,0.67) higher among beneficiaries with lung cancer compared to those without lung cancer; the risk difference increased over time and remained statistically significant at 5 years after diagnosis (RD 1.1%; 95%CI: 0.38,1.8; Figure 3, Table S3). The risk of death was substantially higher among those with lung cancer as compared to those without at 1 year (RD 25.0; 95%CI: 24.6,25.3) and 2 years (RD 33.5; 95%CI: 33.1,33.9). In sensitivity analyses using beneficiaries with COPD as controls, the 5-year risk difference for dementia was attenuated for overall and for most subgroups; dementia risk was higher in controls with COPD for males and beneficiaries diagnosed with lung cancer at age at least 50 (Table S3). The 5-year risk of death was 48.0% (95%CI: 47.4,48.5) among those with lung cancer and compared to 7.1% (95%CI: 6.8, 7.5) among those without lung cancer (RD 40.9%; 95%CI: 40.2,41.6; Table S4). Findings were comparable for beneficiaries less than 50 (5-yr RD:41.2%; 95%CI:40.5%, 41.9%) and in sensitivity analyses with beneficiaries with COPD as controls (5-yr RD:39.6%; 95%CI:38.9%, 40.2%).

**Figure 1.**
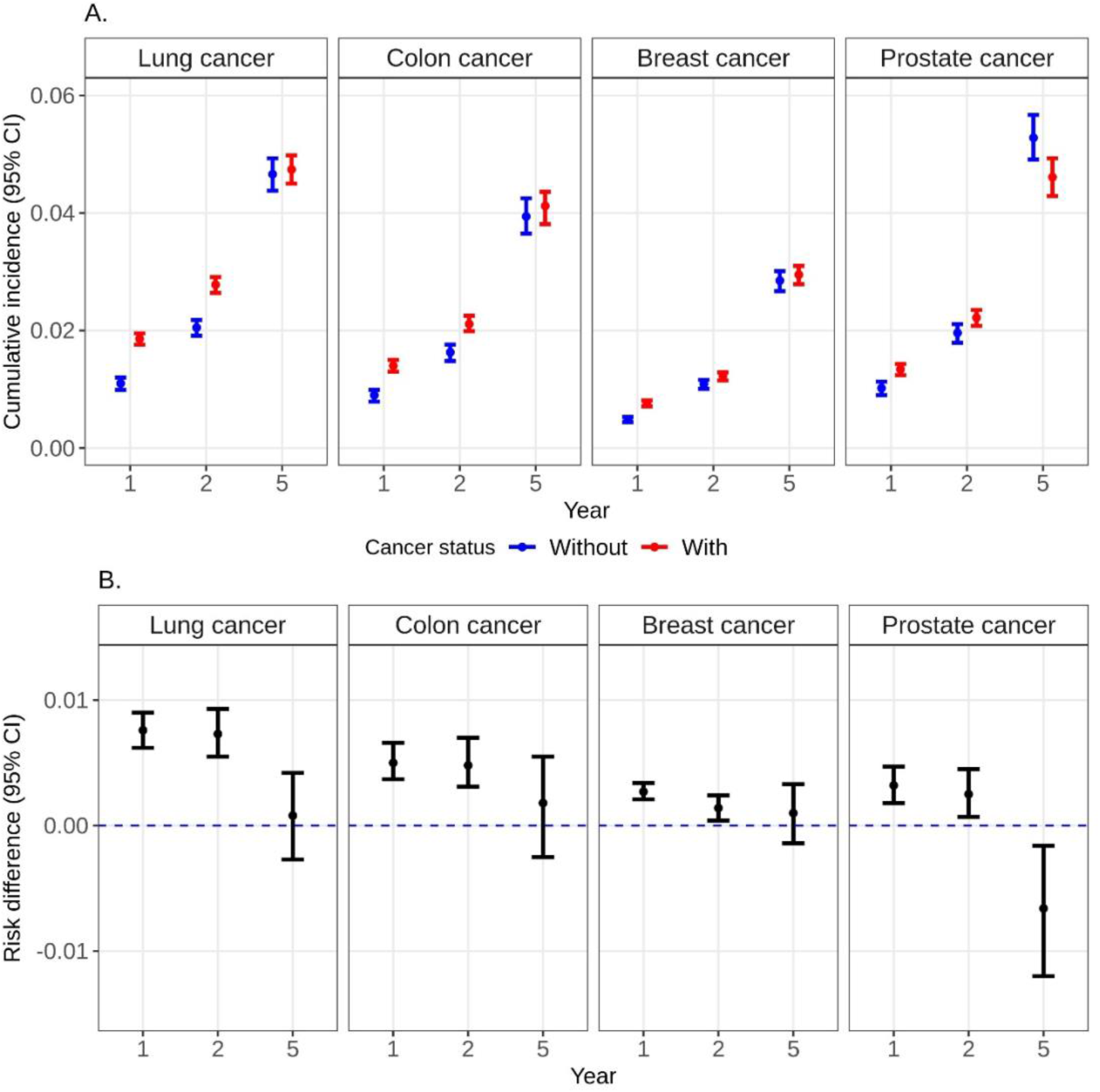
A Weighted cumulative incidence (risk%) of dementia at years 1, 2, and 5 after index date (cancer diagnosis date) among Medicaid beneficiaries matched on cancer status by age, sex, race/ethnicity, year and state. B. Risk difference of dementia at years 1, 2, and 5 after index date (cancer diagnosis date) among Medicaid beneficiaries with HIV matched on cancer status by age, sex, race/ethnicity, year and US state.

**Figure 2.**
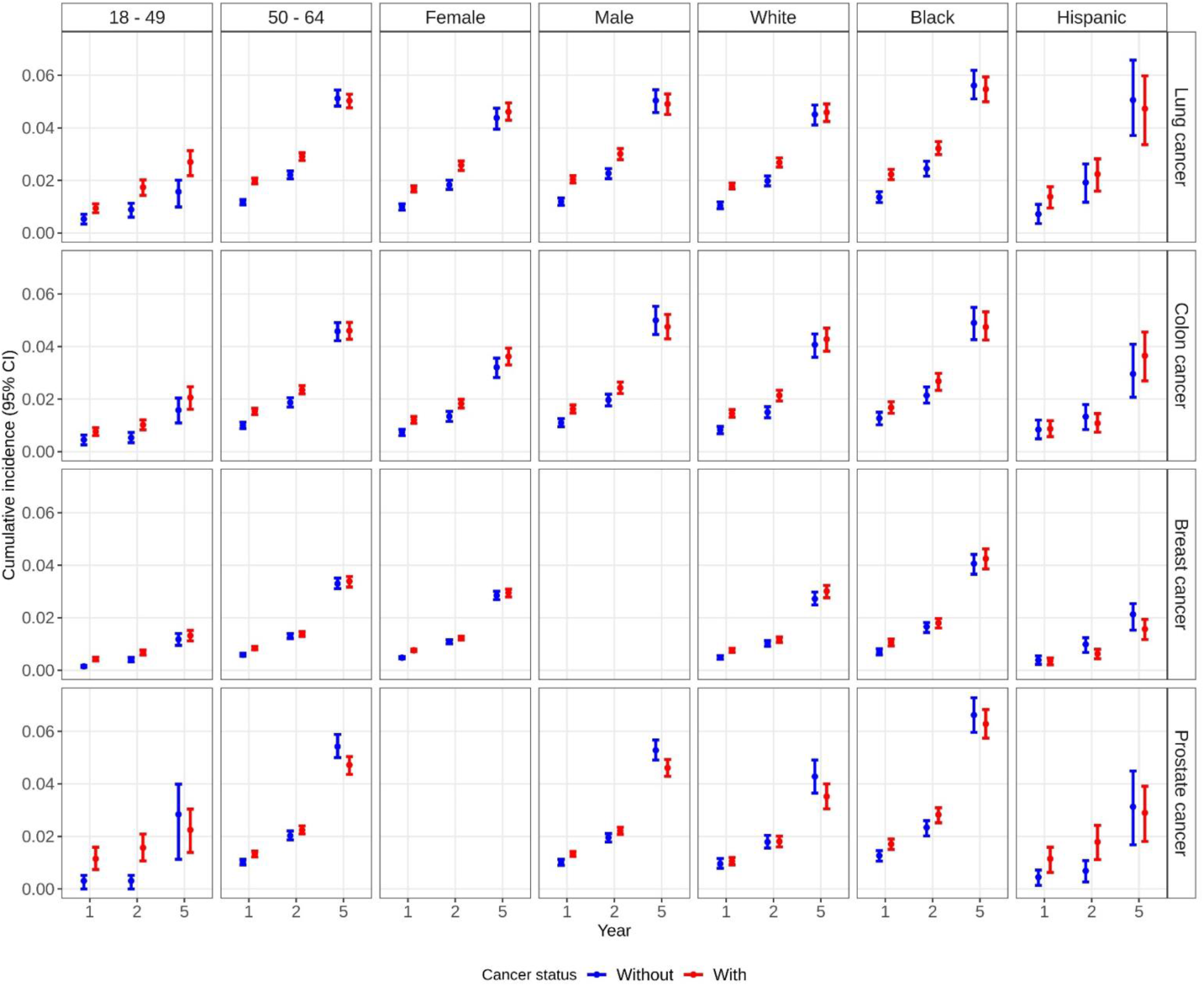
Weighted cumulative incidence (risk%) of dementia at years 1, 2, and 5 after index date (cancer diagnosis date) among Medicaid beneficiaries matched on cancer status by age, sex, race/ethnicity, year and US state, stratified by age at cancer diagnosis, sex, and race/ethnicity.

**Figure 3.**
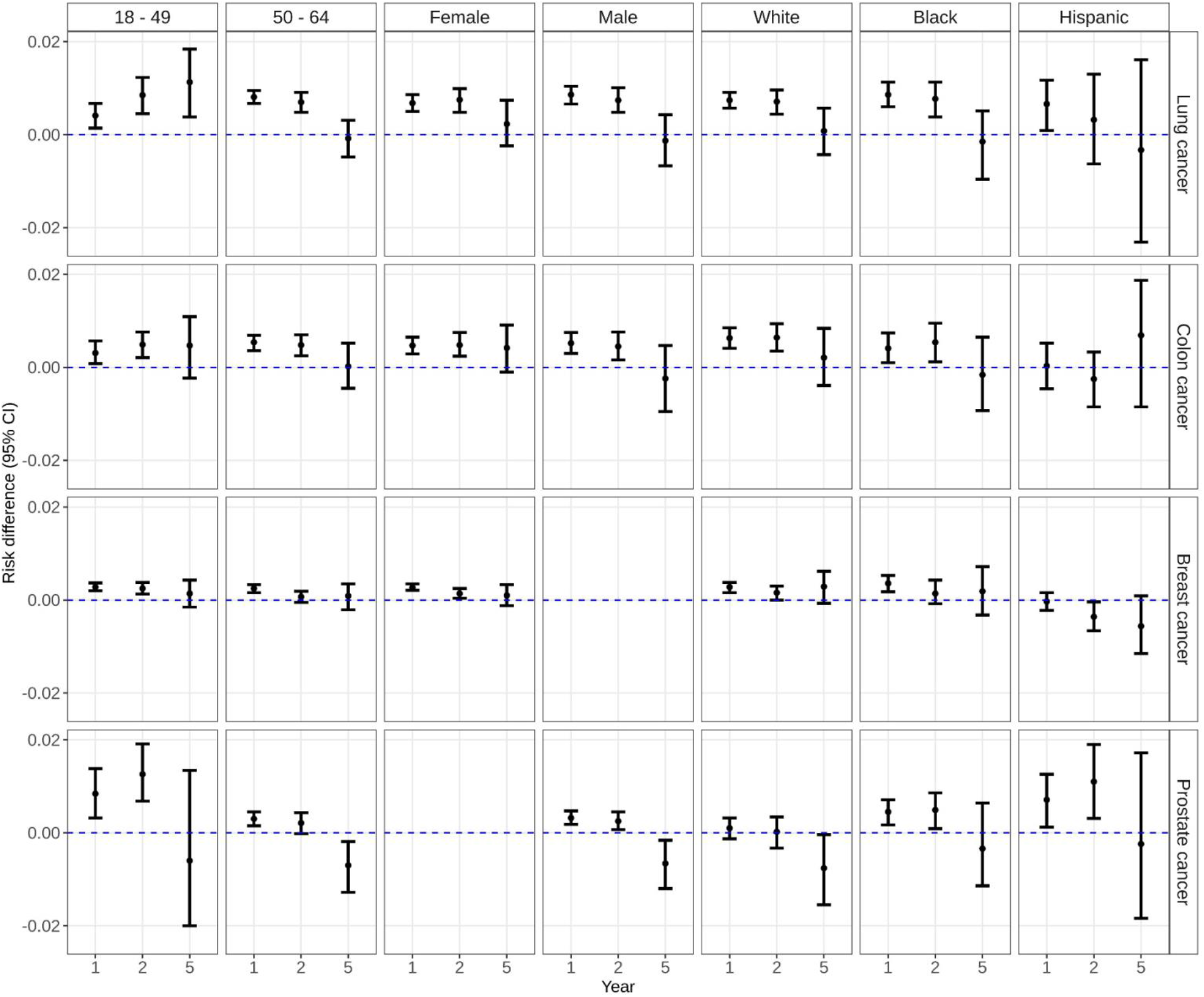
Weighted risk difference of dementia at years 1, 2, and 5 after index date (cancer diagnosis date) among Medicaid beneficiaries matched on cancer status by age, sex, race/ethnicity, year and US state and stratified by age at cancer diagnosis, sex, and race/ethnicity.

### Colon Cancer

The 1-yr risk of dementia was 1.4% (95%CI: 1.3,1.5) among those with colon cancer as compared to 0.90% (95%CI: 0.79,0.99) among those without colon cancer (Figure 1, Table S3). The 5-yr risk of dementia rose to 4.1% (95%CI: 3.8, 4.4) and 3.9% (95%CI:3.7,4.3) among those with and without colon cancer, respectively. There was a 0.50% (95%CI: 0.37,0.66) higher risk of dementia among beneficiaries with colon cancer compared to those without colon cancer at 1 year; findings were similar at 2 years and attenuated at 5 years (RD 0.18; 95%CI: -0.25,0.55). This pattern was similar when beneficiaries were stratified by age, sex, and race/ethnicity groups (Figure 2, Figure 3, Table S3). By contrast, in sensitivity analyses using beneficiaries with comorbidities as controls, the risk difference for dementia after 5 years was modestly higher in controls with comorbidities overall (5-yr RD:-0.9%; 95%CI: -1.3,-0.5) and for all subgroups except those aged younger than 50 and Hispanic beneficiaries (Table S3). The 5-year risk of death was 21.3% (95%CI: 20.8,21.8) among those with colon cancer and compared to 5.2% (95%CI: 4.8, 5.5) among those without colon cancer (RD 16.1%; 95%CI: 15.5,16.8; Table S4). Findings were comparable in sensitivity analyses with beneficiaries with comorbidities as controls (5-yr RD:14.9%; 95%CI:14.3%, 15.6%).

### Breast Cancer

The 1-year risk of dementia was 0.76% (95%CI: 0.71,0.81) among those with breast cancer within 1 year of diagnosis compared to 0.48% (95%CI: 0.44,0.53) among those without breast cancer (Figure 1, Table S3). The 5-year risk of dementia rose to 3.0% (95%CI: 2.8,3.1) and 2.9% (95%CI: 2.7,3.0) among those with and without breast cancer, respectively. There was a 0.27% (95%CI: 0.21,0.34) higher risk of dementia among beneficiaries with breast cancer compared to those without breast cancer at 1 year; findings were attenuated at 5 years (RD 0.10; 95%CI: -0.14,0.43). This pattern was similar when beneficiaries were stratified by age and race/ethnicity groups (Figure 2, Figure 3, Table S3). The 5-year risk of death was 8.8% (95%CI: 8.6,9.1) among those with breast cancer and compared to 3.0% (95%CI: 2.8, 3.2) among those without breast cancer (RD 5.9%; 95%CI: 5.6,6.1; Table S4).

### Prostate Cancer

The 1-year risk of dementia was 1.3% (95%CI: 1.2,1.4) among those with prostate cancer compared to 1.0% (95%CI: 0.9,1.1) among those without prostate cancer (Figure 1, Table S3). The 5-year risk of dementia was 4.6% (95%CI: 4.3,4.9) and 5.3% (95%CI: 4.9,5.7) among those with and without prostate cancer, respectively. There was a 0.32% (95%CI: 0.18,0.47) higher risk of dementia among beneficiaries with prostate cancer compared to those without lung cancer at 1 year; findings were similar at 2 years but inverse at 5 years (RD -0.66; 95%CI: -1.2,-0.16). This pattern was similar when beneficiaries were stratified by age and race/ethnicity groups (Figure 2, Figure 3, Table S3). The 5-year risk of death was 9.8% (95%CI: 9.3,10.3) among those with prostate cancer and compared to 6.8% (95%CI: 6.4, 7.2) among those without prostate cancer (RD 3.0%; 95%CI: 2.3,3.6; Table S4).

## Discussion

In this matched study of Medicaid beneficiaries, the risk of early-onset dementia (EOD) peaked at approximately 4-5% among beneficiaries with and without cancer. While we found modest increases in the risk of early-onset dementia (EOD) in the first 1-2 years after diagnosis of lung, colon, breast, and prostate cancers, these risk differences were less than 1% and attenuated by 5 years post-diagnosis with two exceptions. Beneficiaries diagnosed with lung cancer at age 50 or younger still had a significantly higher risk of EOD than their cancer free counterparts at 5 years post-diagnosis; this finding was attenuated when beneficiaries with lung cancer were compared to beneficiaries with COPD. In contrast, men with prostate cancer, particularly those diagnosed at age of 50 or over, had a significantly decreased risk of EOD as compared to their cancer-free counterparts. These findings do not support an association between cancer and EOD within 5 years of diagnosis.

We found a relatively high proportion of Medicaid beneficiaries, approximately 4% within 5 years, received a diagnosis of EOD. These findings are comparable to recent analyses of Medicare beneficiaries aged 45-65 which reported age-standardized prevalences of 3% or higher (15, 16). Notably, Medicare provides coverage for beneficiaries below the age of 65 if they have a qualifying disability or end-stage renal disease (17). Hence, this population is enriched for comorbid conditions. Similarly, our study included Medicaid beneficiaries, who have a higher comorbidity burden than privately insured adults (18), with and without cancer. General population studies have suggested EOD to range between 8.3 to 22.8 cases per 100,000 person-years for those between the ages of 45-64 (19). These results highlight the burden of EOD, particularly among middle-aged populations with high comorbidity burden.

Recent meta-analyses of cancer-specific studies and all-cause dementia or Alzheimer Disease reported inverse or no association for lung, colorectal, breast, and prostate cancers (11, 12). Both analyses noted heterogeneity in findings for most cancers. In our study, we observed a modestly increased risk of EOD within 1-2 years of diagnosis for lung, colon, breast, and prostate cancers that generally attenuated or became inverse (prostate) by 5 years. While our findings at 5 years post diagnosis are generally compatible with prior studies for average-onset dementia (11, 12), the positive association within 2 years of diagnosis is not. Given this pattern of association was consistent across all cancer types, which encompass varying risk factors, treatment types, and prognosis, it is likely that detection bias contributes to the positive findings. It is possible that dementia is more likely to be diagnosed among adults under medical surveillance for other conditions like cancer, particularly among those below the age of 65. This is further supported by the attenuation of the association by 5 years post-diagnosis at which point many beneficiaries surviving their cancer may be in a less intensive surveillance phase of treatment (20).

In contrast to this general pattern, beneficiaries diagnosed with lung cancer at age 49 or younger maintained a significantly higher risk of EOD than their cancer free counterparts at 5 years post-diagnosis. To better account for differences in smoking status among matched pairs for lung cancer, we conducted sensitivity analyses in which lung cancer-free matches were selected from beneficiaries with a diagnosis of chronic pulmonary disease. In this analysis, the finding between lung cancer and dementia at 5 years post diagnosis was attenuated when beneficiaries with lung cancer were compared to beneficiaries with chronic pulmonary disease. This attenuation was largely due to an increase in EOD among cancer-free controls. While our analyses did adjust for co-morbidity burden, this finding may suggest that shared risk factors for lung cancer and COPD, like smoking, may be more important to EOD risk than the lung cancer diagnosis/treatment itself.

We evaluated the cumulative incidence of dementia among beneficiaries with and without cancer who were enrolled in Medicaid in 26 states over a 20-year period. In our study, beneficiaries who were diagnosed with cancer were similar to those without cancer with respect to demographic factors due to matching. However, differences in unmeasured factors, such as smoking status, could influence both cancer and dementia incidence. When possible, we conducted sensitivity analyses to compare beneficiaries with cancer to cancer-free controls with conditions that could influence both cancer and dementia, including beneficiaries with smoking-associated conditions for lung cancer and beneficiaries with metabolic conditions for colon cancer. The size of our population allowed us to evaluate outcomes by age and assess sex-specific cancers. However, we were unable to assess differences in cancer stage at diagnosis, which could reflect differences in survival and treatment that could impact dementia risk. Given profound bias concerns that have been highlighted in analyses of cancer and dementia, we designed our analyses to minimize these concerns as much as possible. To address potential survival bias, we conducted analyses in which beneficiaries were matched on time at risk and assessed dementia incidence as clearly specified time points after cancer diagnosis – 1, 2, or 5 years. We also accounted for the competing risk of death in all analyses. However, we cannot rule out the potential for detection bias among beneficiaries with cancer, though sensitivity analyses among those with other chronic conditions may minimize this concern.

We investigated the association between cancer and early-onset dementia among Medicaid beneficiaries. The incidence of EOD in our study population was comparable to those reported in other studies US populations with high comorbidity burden (15, 16) and much higher than general population studies in several developed countries (19), which further supports the need for additional work into methods to prevent, detect, and treat this condition. We observed early, modest increases in EOD after diagnosis of colon, breast and prostate cancer that attenuated or became inverse by 5 years post diagnosis. Diagnosis with lung cancer, particularly below the age of 50, was associated with a modestly increased risk of EOD 5 years after diagnosis; though this positive association may be due to shared risk factors, like smoking. Our findings for EOD are compatible with prior studies of cancer and average-onset dementia and suggest that cancer diagnosis is not a strong risk factor for early-onset dementia even among low SES population enriched for other risk factors.

## Data Availability

The data that support the findings of this study are under the authority of the Centers for Medicare & Medicaid Services (CMS) and administered by ResDAC. Investigators may reuse these data if they independently meet CMS requirements and obtain both CMS reuse approval and permission from the studys NIH program officer.

