## Supplemental Tables and Figures for "Assessing the risk of early-onset dementia within 5 years of cancer diagnosis"

### Supplementary Methods

We matched beneficiaries by cancer status for incident diagnoses of lung, breast, colon, and prostate cancers. We used incidence density matching with replacement to match beneficiaries with cancer to beneficiaries without cancer at the date of cancer diagnosis. Beneficiaries with cancer diagnosis after $b_0$ (the exposed) were sorted ascendingly by date of diagnosis (Figure S1). Using the sorted data, the matching pool for exposed beneficiary *i* was composed of all the unexposed at the time of cancer diagnosis $t_i$ (index date), regardless of future exposure. Any unexposed in the matching pool who had a dementia diagnosis, died, turned 65 years old, or ended Medicaid enrollment before $t_i$ was removed. The beneficiaries in the matching pool were matched to the exposed by age at $b_0$ +/- 2 years, sex (lung and colon cancers), race/ethnicity, state, and year at $b_0$ +/- 2 years. Each exposed and all its matches formed a cluster. For each cluster *i*, the index date, $t_i$, was set as the new analytic baseline. Matches with any cancer or dementia diagnosis prior to $t_i$ were excluded.

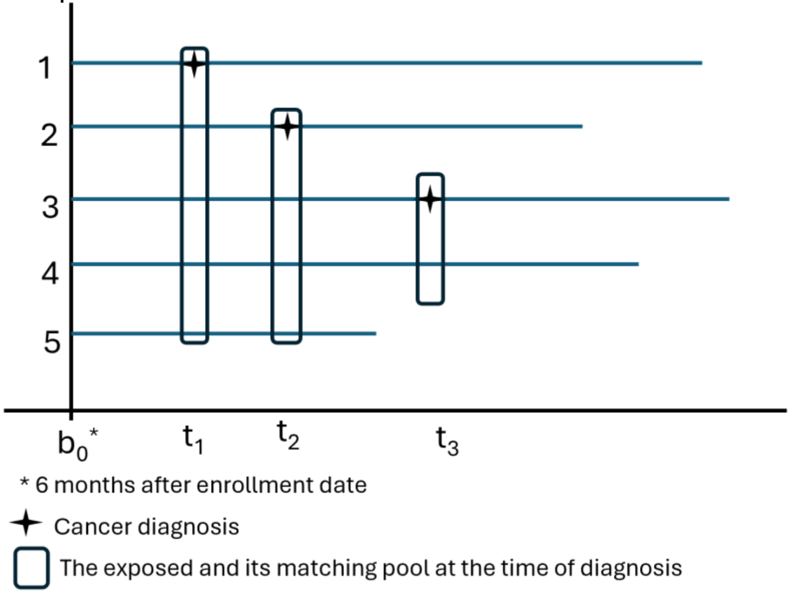


Figure S1. Diagram illustrating the steps on how the matching pool for the exposed group were obtained.

### Supplementary tables and figures

Table S1. List of ICD codes used in the study

| Variable | ICD-9 Code[a] | ICD-10 Code[a] |
|---|---|---|
| **Cancers** | | |
| Lung | 162.2-162.5, 162.8, 162.9 | C34.x |
| Colon | 153.x | C18.x |
| Breast | 174.x, 175.x | C50.x |
| Prostate | 185.x | C61.x |
| Any cancer | 140.x-195.x, 199.x-208.x | C00.x-C43.x, C45.x-C76.x, C80.x-C96.x |
| **Dementia** | 046.19, 290.4x, 294.0, 294.1x, 294.2x, 294.8, 294.9, 331.0, 331.1x, 331.5, 331.6, 331.7, 331.8x, 331.82, 438.0, 781.8 | A81.00, A81.09, F01.x, F02.8x, F03.9x, F04, G30.x, G31.0x, G31.83, G31.85, G91.2, G94, I69.x, R41.4 |
| **Charlson comorbidities** | | |
| Myocardial infarction | 410.x, 412.x | I21.x, I22.x, I25.2 |
| Congestive heart failure | 398.91, 402.01, 402.11, 402.91, 404.01, 404.03, 404.11, 404.13, 404.91, 404.93, 425.4-425.9, 428.x | I09.9, I11.0, I13.0, I13.2, I25.5, I42.0, I42.5-I42.9, I43.x, I50.x, P29.0 |
| Peripheral vascular disease | 093.0, 437.3, 440.x, 441.x, 443.1-443.9, 447.1, 557.1, 557.9, V43.4 | I70.x, I71.x, I73.1, I73.8, I73.9, I77.1, I79.0, I79.2, K55.1, K55.8, K55.9, Z95.8, Z95.9 |
| Cerebrovascular disease | 362.34, 430.x-438.x | G45.x, G46.x, H34.0, I60.x-I69.x |
| Chronic pulmonary disease | 416.8, 416.9, 490.x-505.x, 506.4, 508.1, 508.8 | I27.8, I27.9, J40.x-J47.x, J60.x-J67.x, J68.4, J70.1, J70.3 |
| Rheumatic disease | 446.5, 710.0-710.4, 714.0-714.2, 714.8, 725.x | M05.x, M06.x, M31.5, M32.x-M34.x, M35.1, M35.3, M36.0 |
| Peptic ulcer disease | 531.x-534.x | K25.x-K28.x |
| Mild liver disease | 070.22, 070.23, 070.32, 070.33, 070.44, 070.54, 070.6, 070.9, 570.x, 571.x, 573.3, 573.4, 573.8, 573.9, V42.7 | B18.x, K70.0-K70.3, K70.9, K71.3-K71.5, K71.7, K73.x, K74.x, K76.0, K76.2-K76.4, K76.8, K76.9, Z94.4 |
| Diabetes without chronic complication | 250.0-250.3, 250.8, 250.9 | E10.0, E10.l, E10.6, E10.8, E10.9, E11.0, E11.1, E11.6, E11.8, E11.9, E12.0, E12.1, E12.6, E12.8, E12.9, E13.0, E13.1, E13.6, E13.8, E13.9, E14.0, E14.1, E14.6, E14.8, E14.9 |

| | | |
|---|---|---|
| Diabetes with chronic complication | 250.4-250.7 | E10.2-E10.5, E10.7, E11.2-E11.5, E11.7, E12.2-E12.5, E12.7, E13.2-E13.5, E13.7, E14.2-E14.5, E14.7 |
| Hemiplegia or paraplegia | 334.1, 342.x, 343.x, 344.0-344.6, 344.9 | G04.1, G11.4, G80.1, G80.2, G81.x, G82.x, G83.0-G83.4, G83.9 |
| Renal disease | 403.01, 403.11, 403.91, 404.02, 404.03, 404.12, 404.13, 404.92, 404.93, 582.x, 583.0-583.7, 585.x, 586.x, 588.0, V42.0, V45.1, V56.x | I12.0, I13.1, N03.2-N03.7, N05.2-N05.7, N18.x, N19.x, N25.0, Z49.0-Z49.2, Z94.0, Z99.2 |
| Moderate or severe liver disease | 456.0-456.2, 572.2-572.8 | I85.0, I85.9, I86.4, I98.2, K70.4, K71.1, K72.1, K72.9, K76.5, K76.6, K76.7 |
| HIV | 042, 079.53, 795.71, V08 | B20, B97.35, R75, Z21 |

[a] A code ending with ".x" indicates a wildcard. Any number could appear after the decimal place.

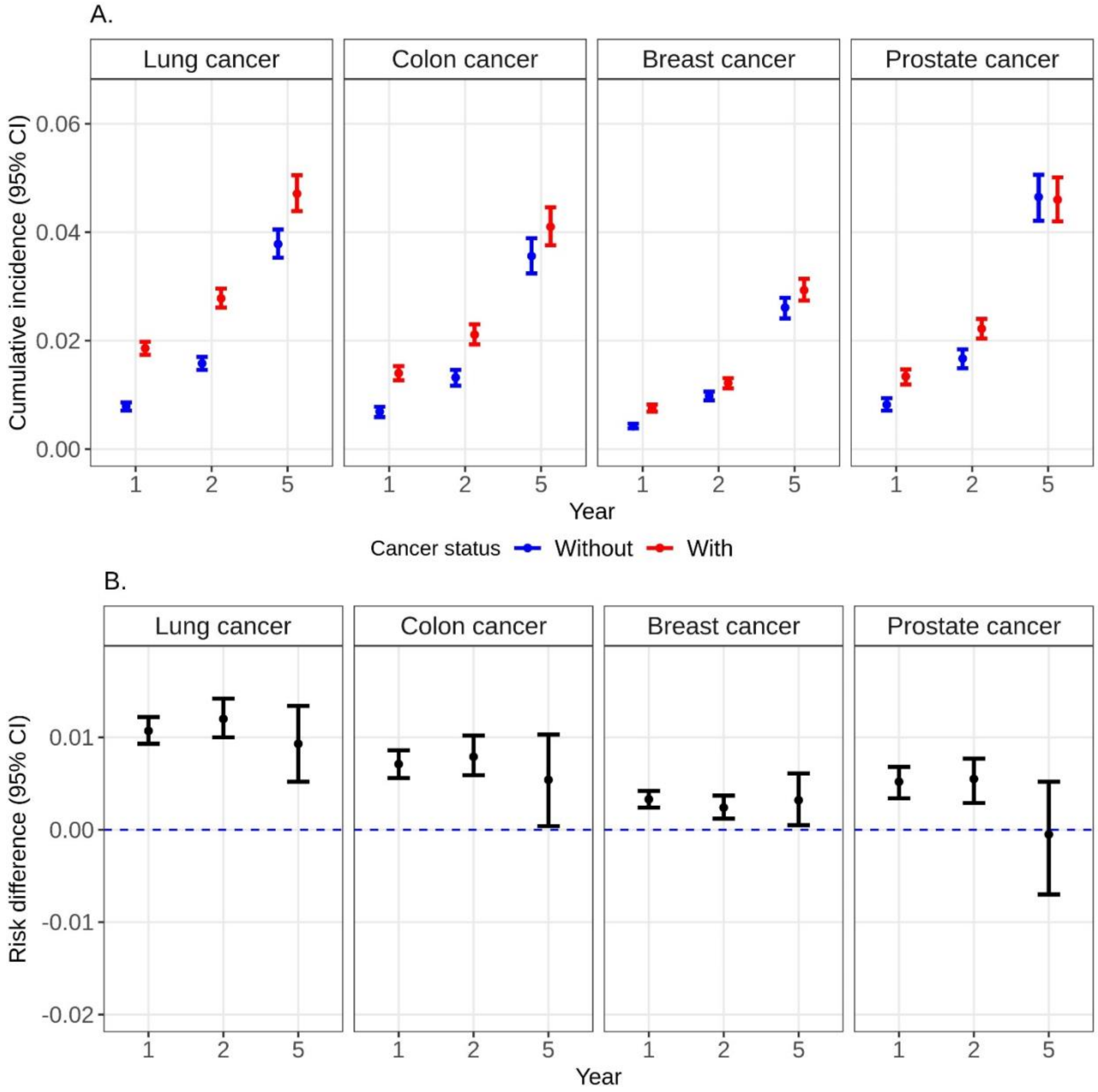


Figure S2. A. Crude cumulative incidence (risk%) and B. crude risk difference of dementia at years 1, 2, and 5 after baseline among matched Medicaid beneficiaries by cancer status.

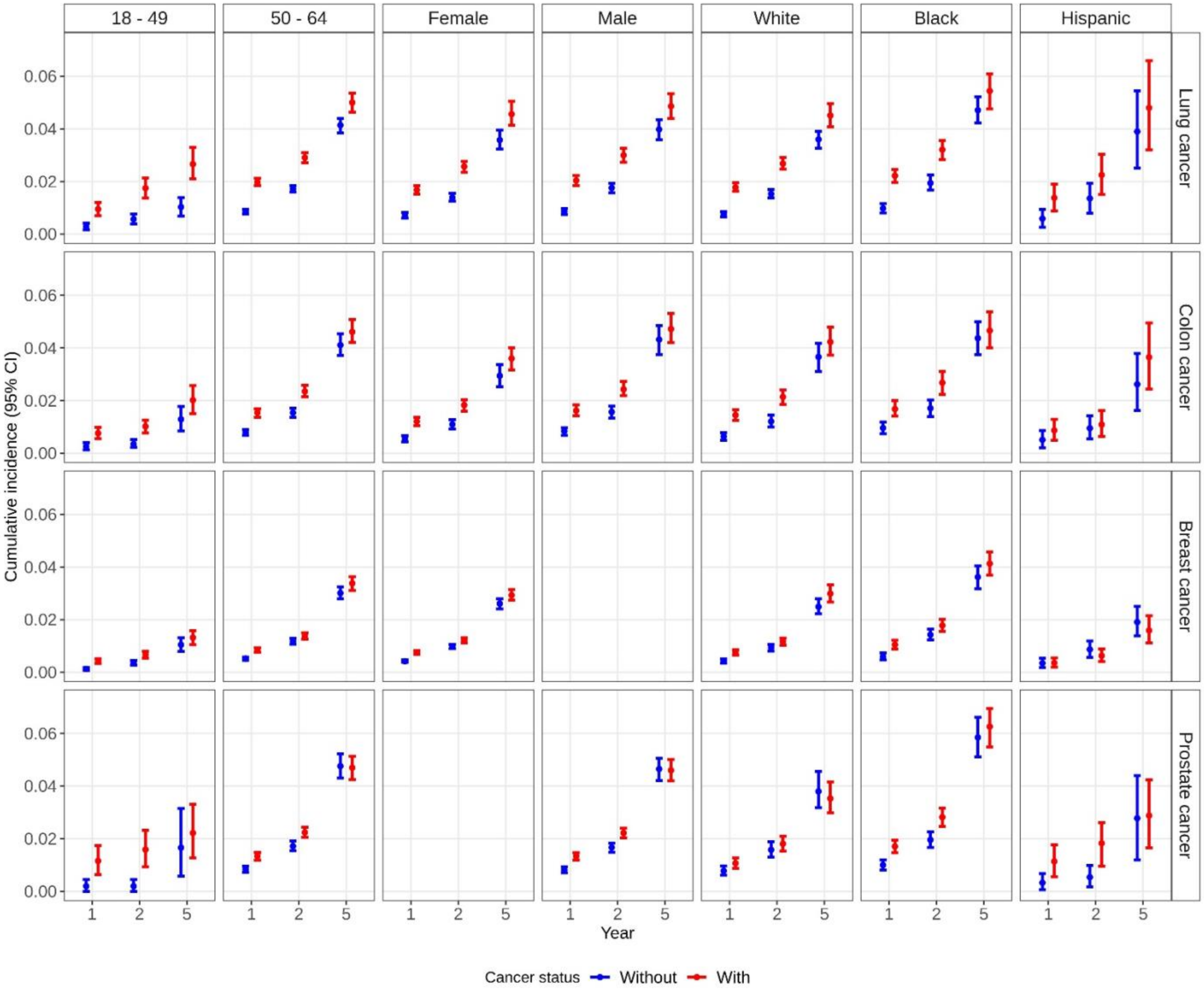


Figure S3. Crude cumulative incidence (risk%) of dementia at years 1, 2, and 5 after baseline among matched Medicaid beneficiaries by cancer status, age, sex, and race/ethnicity.

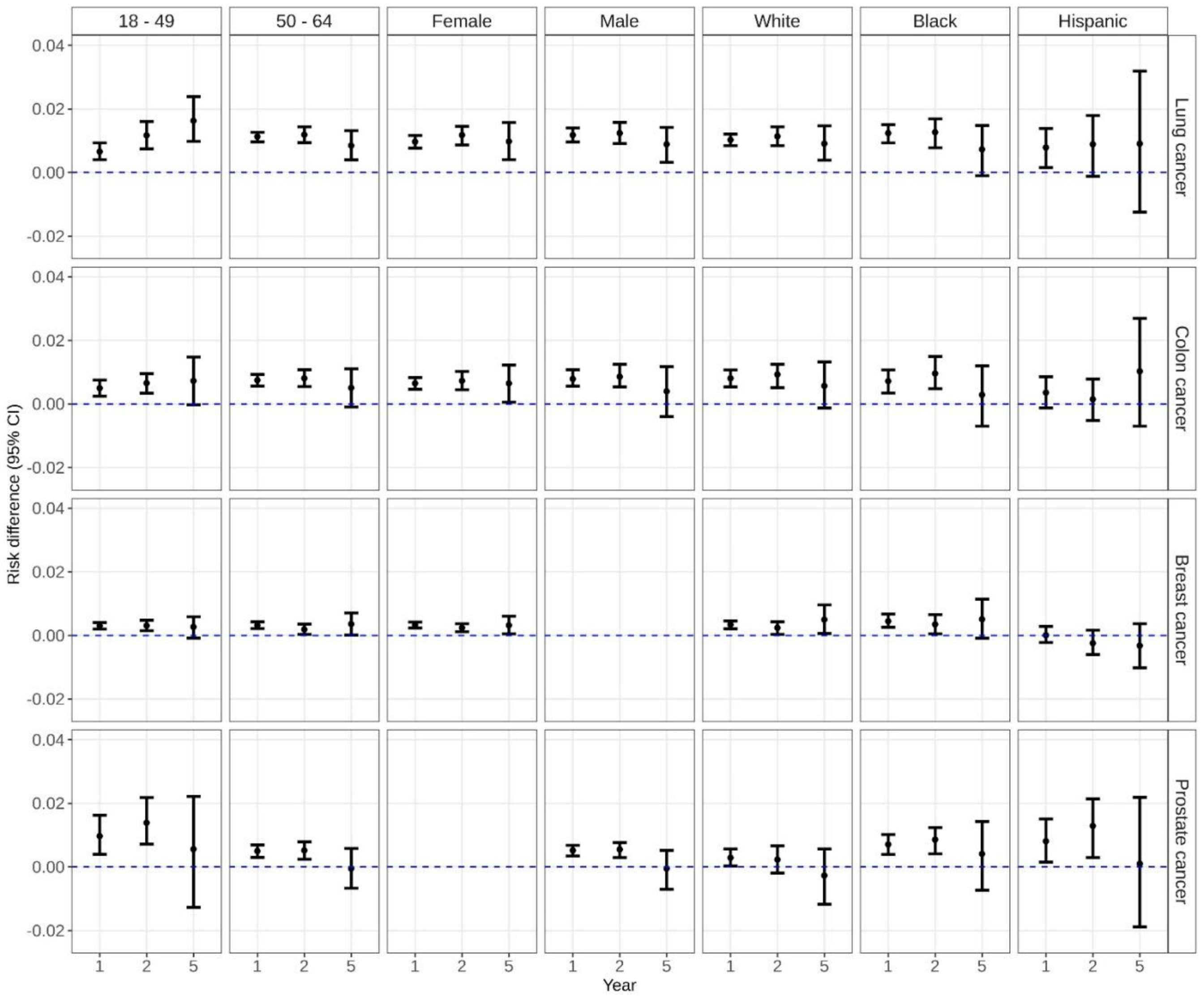


Figure S4. Crude risk difference of dementia at years 1, 2, and 5 after baseline among matched Medicaid beneficiaries by cancer status, age, sex, and race/ethnicity.

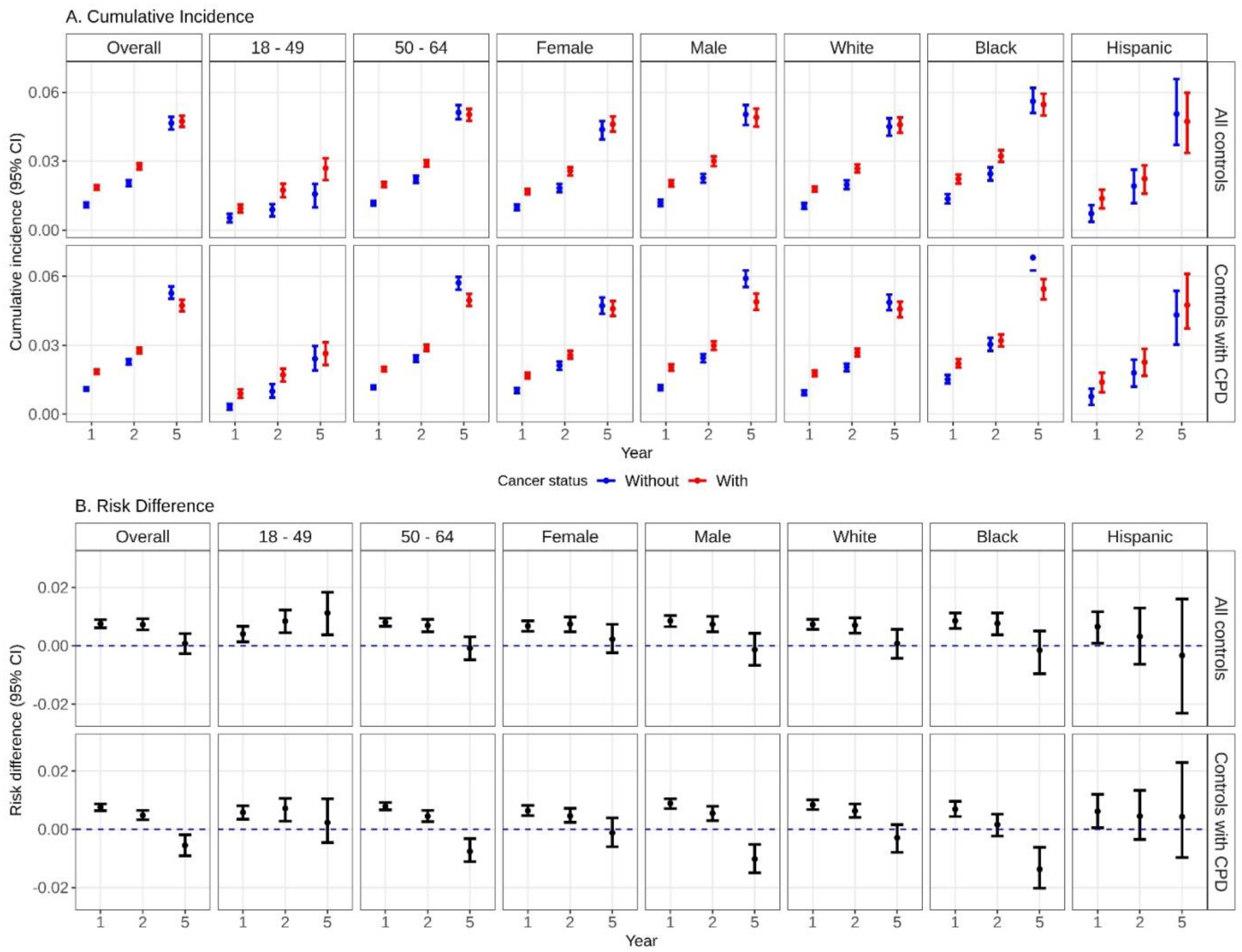


Figure S5. A. Weighted cumulative incidence (risk%) and B. weighted risk difference of dementia at years 1, 2, and 5 after baseline comparing Medicaid beneficiaries with lung cancer and their matched controls (all controls – 1st row of each panel, controls with chronic pulmonary disease – 2nd row of each panel), overall and stratified by age, sex, and race/ethnicity.

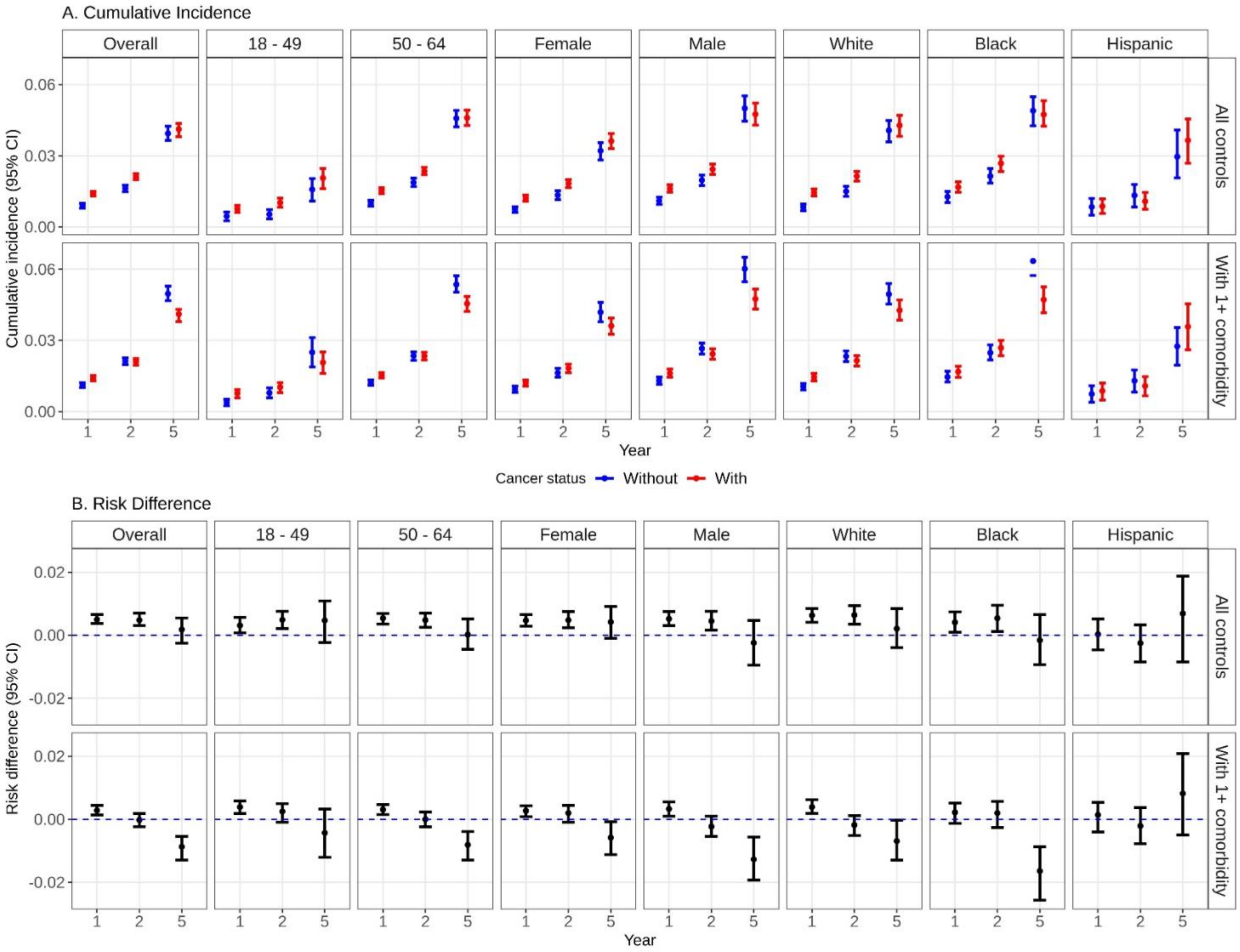


Figure S6. A. Weighted cumulative incidence (risk%) and B. weighted risk difference of dementia at years 1, 2, and 5 after baseline comparing Medicaid beneficiaries with colon cancer and their matched controls (all controls – 1st row of each panel, controls with ≥1 comorbidity – 2nd row of each panel), overall and stratified by age, sex, and race/ethnicity.

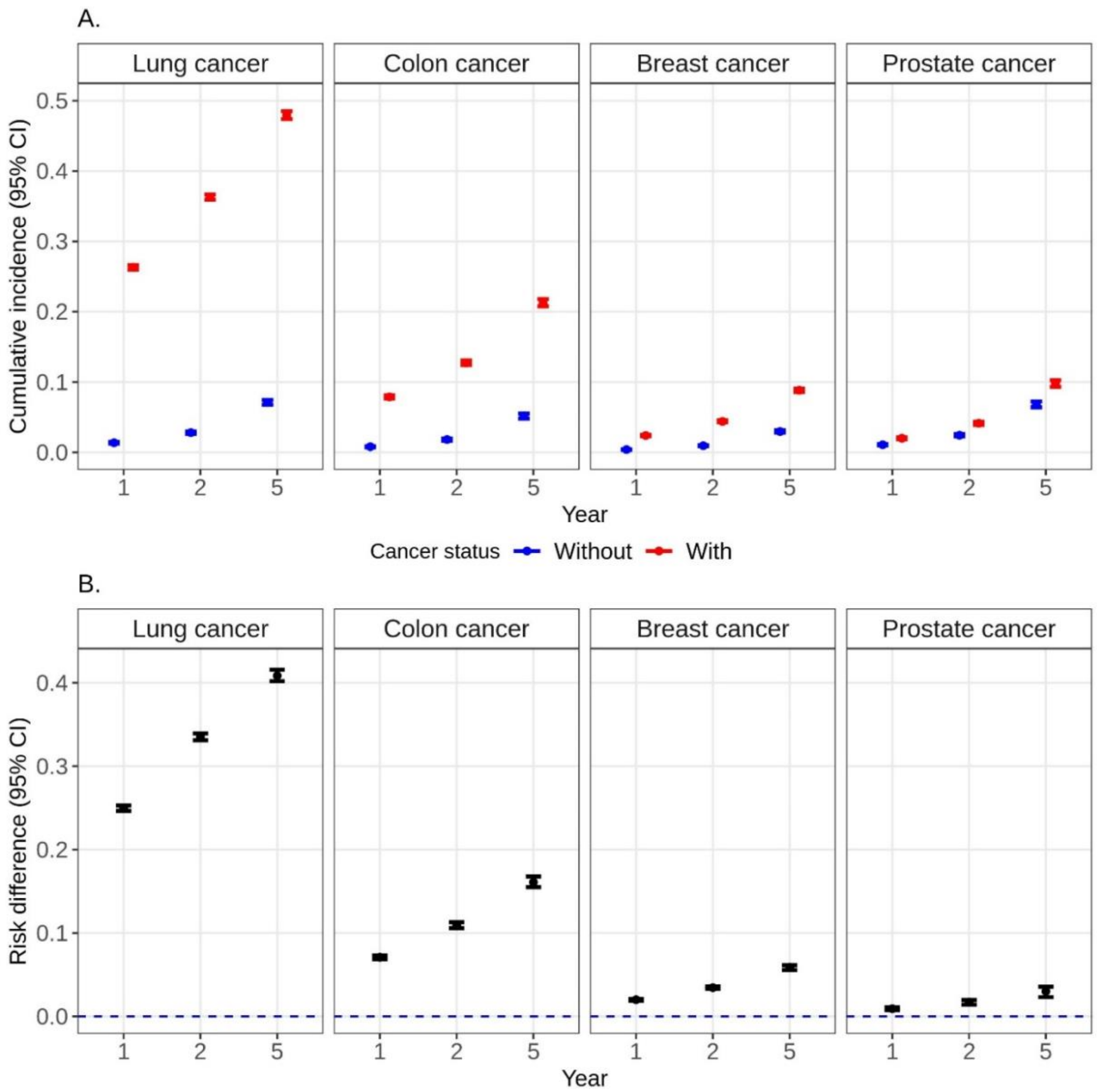


Figure S7. A. Weighted cumulative incidence (risk%) and B. weighted risk difference of death at years 1, 2, and 5 after baseline among matched Medicaid beneficiaries by cancer status.

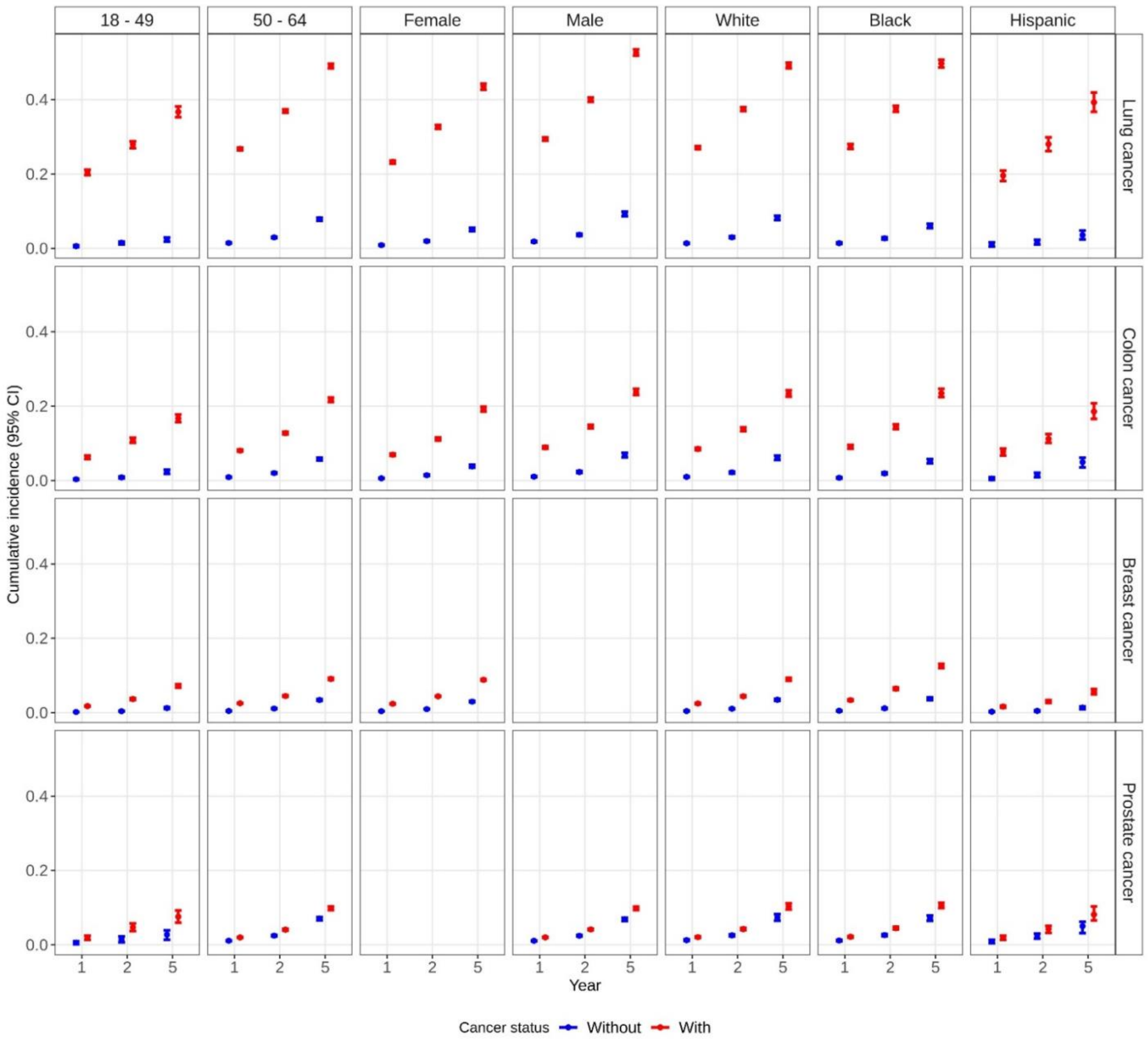


Figure S8. Weighted cumulative incidence (risk%) of death at years 1, 2, and 5 after baseline among matched Medicaid beneficiaries by cancer status, age, sex, and race/ethnicity.

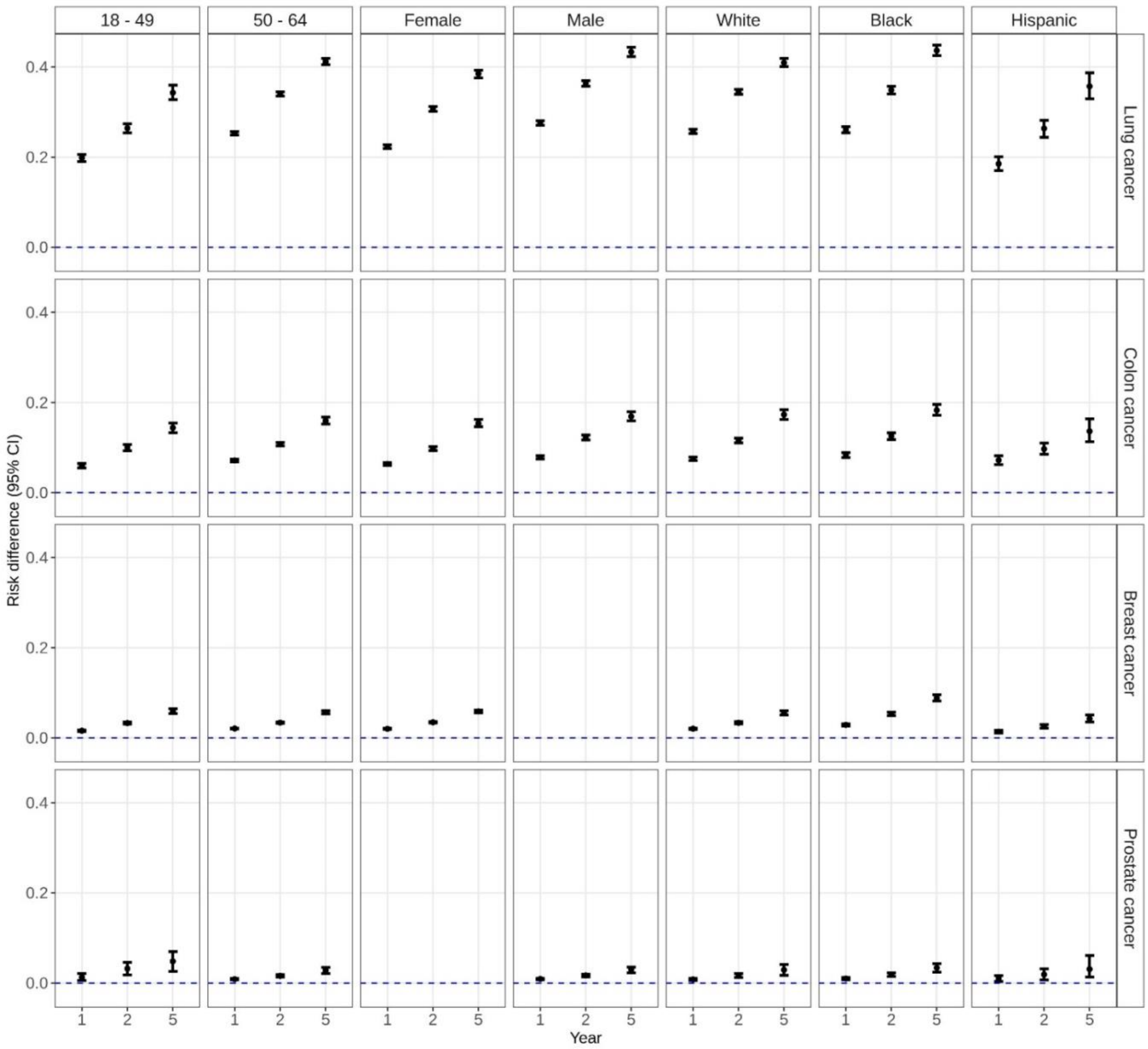


Figure S9. Weighted risk difference of death at years 1, 2, and 5 after baseline among matched Medicaid beneficiaries by cancer status, age, sex, and race/ethnicity.
